# DO NOT LOSE SLEEP OVER IT: IMPLANTED BRAIN-COMPUTER INTERFACE FUNCTIONALITY DURING NIGHTTIME IN LATE-STAGE AMYOTROPHIC LATERAL SCLEROSIS

**DOI:** 10.1101/2024.10.11.24315027

**Authors:** Sacha Leinders, Erik J. Aarnoutse, Mariana P. Branco, Zac V. Freudenburg, Simon H. Geukes, Anouck Schippers, Malinda S.W. Verberne, Max van den Boom, Benny van der Vijgh, Nathan E. Crone, Timothy Denison, Nick F. Ramsey, Mariska J. Vansteensel

## Abstract

**Background and objectives:** Brain-computer interfaces (*BCIs*) hold promise as augmentative and alternative communication technology for people with severe motor and speech impairment (locked-in syndrome) due to neural disease or injury. Although such BCIs should be available 24/7, to enable communication at all times, feasibility of nocturnal BCI use has not been investigated. Here, we addressed this question using data from an individual with amyotrophic lateral sclerosis (ALS) who was implanted with an electrocorticography-based BCI that enabled the generation of click-commands for spelling words and call-caregiver signals.

**Methods:** We investigated nocturnal dynamics of neural signal features used for BCI control, namely low (*LFB*: 10–30Hz) and high frequency band power (*HFB*: 65-95Hz). Additionally, we assessed the nocturnal performance of a BCI decoder that was trained on daytime data by quantifying the number of unintentional BCI activations at night. Finally, we developed and implemented a nightmode decoder that allowed the participant to call a caregiver at night, and assessed its performance.

**Results:** Power and variance in HFB and LFB were significantly higher at night than during the day in the majority of the nights, with HFB variance being higher in 88% of nights. Daytime decoders caused 245 unintended selection-clicks and 13 unintended caregiver-calls per hour when applied to night data. The developed nightmode decoder functioned error-free in 79% of nights over a period of ±1.5 years, allowing the user to reliably call the caregiver, with unintended activations occurring only once every 12 nights.

**Discussion:** Reliable nighttime use of a BCI requires decoders that are adjusted to sleep-related signal changes. This demonstration of a reliable BCI nightmode and its long-term use by an individual with advanced ALS underscores the importance of 24/7 BCI reliability.

**Trial registration:** This trial is registered in clinicaltrials.gov under number NCT02224469 (https://clinicaltrials.gov/study/NCT02224469?term=NCT02224469&rank=1). Date of submission to registry: August 21, 2014. Enrollment of first participant: September 7, 2015.

## 1. INTRODUCTION

Neural disease or injury can lead to a state of almost complete paralysis while cognition is spared, so called locked-in syndrome (*LIS*; American Congress of Rehabilitation Medicine, 1995; Smith & Delargy, 2005). The ability to communicate is a determining factor for the quality of life in people with LIS (Corallo et al., 2017; Rousseau et al., 2015). Conventional augmentative and alternative communication (*AAC*) technology can assist people with LIS in communication but requires residual, reliable motor control (Higginbotham et al., 2007), which not all people with LIS are able to produce. These individuals may benefit from brain-computer interface (*BCI*) technology, which allows computer control based on brain signals (Nicolas-Alonso and Gomez-Gil, 2012; Wolpaw et al., 2002; Wolpaw and Wolpaw, 2012).

Several studies have shown that people with severe paralysis can use an implanted BCI for communication (Hochberg et al., 2006; Metzger et al., 2023, 2022; Mitchell et al., 2023; Moses et al., 2021; Oxley et al., 2021; Pandarinath et al., 2017; Stavisky et al., 2018; Vansteensel et al., 2016; Willett et al., 2023, 2021). Moreover, first evidence of the feasibility and user benefit of unsupervised at-home use of implanted BCI systems has now been demonstrated (Oxley et al., 2021; Vansteensel et al., 2016; Vansteensel et al., 2024). Successful at-home use of a communication device by people with LIS, however, involves the ability to call caregivers and communicate needs not just during the day, but also at night. Importantly, research has shown that sleep affects brain activity in several regions (Achermann, 2009; Šušmáková, 2004), including the sensorimotor cortex (De Carli et al., 2016; Ramot et al., 2013), which constitutes the signal source of the vast majority of implanted BCIs.

We here used a unique dataset from a BCI user with late-stage amyotrophic lateral sclerosis (ALS) to investigate how BCI control signals in the sensorimotor cortex change at night. Second, we tested the nocturnal performance of BCI decoders that were optimized for daytime usage by applying them, offline, to night data. Finally, we developed and tested a dedicated nightmode decoding algorithm that allowed the BCI user to reliably generate a call-caregiver signal at night and assessed its performance in daily life settings over a period of ± 1.5 years.

## 2. METHODS

### 2.1 Standard Protocol Approvals, Registrations, and Patient Consents

The study was approved by the Medical Research Ethics Committee of the University Medical Center of Utrecht, the Netherlands. Research was conducted according to the Declaration of Helsinki (2013). The study was a registered clinical trial (*ClinicalTrials.gov: NCT02224469*). The participant gave informed consent using a procedure dedicated to people with severe communication impairments (Vansteensel et al., 2016).

### 2.2 Participant Information

The participant of this study is a woman diagnosed with ALS, who was enrolled in the Utrecht NeuroProsthesis clinical trial in September 2015, when she was in her fifties. At the time, she was quadriplegic and anarthric, and used an eye gaze device and eye blinks for communication. She received chronic tracheostomy invasive ventilation. In November 2015, the participant was implanted with a BCI system, which enabled her to generate click-commands for communication. To produce a click-command, she attempted right hand fingertapping (sequential finger opposition) to generate brief changes in sensorimotor neural activity (Vansteensel et al., 2016). A second decoding algorithm (Leinders et al., 2017) detected a more sustained increase of sensorimotor cortex activity (> 7.6 seconds), generated by more long-term attempted fingertapping, which triggered a call-caregiver signal, an escape from software menus, or a system activation from daytime standby mode, depending on the AAC software context, henceforth referred to as *‘escape’*.

### 2.3 Data Acquisition

The implant consisted of two electrocorticography (ECoG) electrode strips (Resume II ®, Medtronic; off-label use), with 4 electrodes each, placed subdurally over the hand knob region of the left sensorimotor cortex. Two other strips were implanted over the dorsolateral prefrontal cortex (Leinders et al., 2020). In October 2015, one strip from each brain region was connected to an amplifier-transmitter device (‘*implanted device’*; Activa ® PC+s, Medtronic; off-label use), which was implanted subcutaneously under the left clavicle. In September 2020 (week 255 since implantation), the implanted device was replaced, to enable continued use of the system (Vansteensel et al., 2024). During this surgery, both sensorimotor cortex electrode strips were connected to the implanted device. The lead of the previously connected DLPFC strip was capped and left disconnected.

The implanted device wirelessly transmitted either raw potential data at 200Hz (time domain) or frequency-decomposed data at 5Hz (power domain) (Vansteensel et al., 2016) to an external antenna. The power domain mode required significantly less battery power than the time domain mode and was therefore always employed for independent at-home use, as well as for nocturnal measurements. Up to December 2021, the participant used a combination of low frequency band (*LFB*: centered around 20Hz) and high frequency band power (*HFB*: centered around 80Hz) from electrode pair E2E3 for daytime BCI control. As of December 2021, LFB power of E2E3 was combined with HFB power from electrode pair E10E12 to boost BCI performance, after it had gradually decreased (Vansteensel et al., 2024). Since only E2E3 data was used for BCI control at night (see *Nightmode Decoder Development* section) all data presented in this manuscript represents LFB and HFB power domain data from that electrode pair.

All data were recorded at the participant’s residence, either during research sessions or during independent at-home use. Night data and research session data that were used to develop decoding algorithms for night usage were recorded with the device that was implanted in 2015. All at-home BCI use data presented in this manuscript were recorded with the new device, between September 2020 and October 2022. The term ‘*recording file*’ refers to an individual data file (created when the BCI at-home use software was started and appended with neural data until the software was closed).

### 2.4 Data Selection Procedure

Based on information from the participant’s caregivers regarding her daily schedule, we assigned files recorded between 12:00h and 20:00h to ‘day data’ and 0:00h and 8:00h to ‘night data’. Data recorded in the other periods were not analyzed because they typically involved extensive physical care. For initial exploration of day and night signals, we only considered days and subsequent nights with at least 6 hours of data (not necessarily continuous recording files). For statistical comparison of day and night neural signal features, we considered daytime and nighttime recording files with a minimum duration of 1 hour and grouped these into 4-week periods, to prevent any longitudinal changes in power from affecting the analyses. For calculating the nocturnal performance of the daytime BCI decoders, we used data from initial night measurements conducted before device replacement. An overview of the different datasets, their use, and the number of datasets is presented in Table 1.

**Table 1:** Overview of different datasets. Datasets are presented in the same order as in the main text. Data recorded before week 255 was recorded with the first implanted device. Week numbers indicate weeks since implantation of the ECoG electrodes. See also Supplementary Figure S1 for a distribution of available data over the 4-week periods used for statistical analysis (4-week datasets).

| **Dataset type:** | **Recorded during:** | **Number of datasets:** | **Used for:** | **Period:** | **Methods Section:** |
| --- | --- | --- | --- | --- | --- |
| Day-night datasets: night data > 6 hours duration with matching daytime dataset | At-home use* | 209 day-night datasets | Initial exploration of day and night signals | Week 255-361 | Data Analysis: Comparing Day and Night ECoG Signals |
| 4-week datasets: Recording files > 1 hour duration, grouped into 4-week periods (see also Supp. Fig. S1) | At-home use* | 24 4-week periods (comprising 499 days and 449 nights with recording files of > 1 hour duration) | Statistical comparison of day and night signals | Week 255-361 | Data Analysis: Comparing Day and Night ECoG Signals |
| Initial night measurements | Passive, at-home data recordings** | 17 nights | Quantification of nocturnal performance of decoders optimized for daytime use; Optimization of nightmode decoder | Week 111-220 | Data Analysis: Performance of Daytime BCI Decoder at Night |
| Research session data | Research sessions | 14 repetitions of the research task (see the respective section) | Optimization of nightmode decoder | Week 220-244 | Nightmode Decoder Development |
| *All data marked with ‘at-home use’ was recorded during periods of active BCI usage. **The initial night measurements were passive night recordings, during which the participant did not attempt to use the BCI. |  |  |  |  |  |

### 2.5 Data Pre-processing Pipeline

ECoG data was analyzed with MATLAB (The Mathworks, Inc., version 2021a). LFB and HFB power data, sampled every 200ms (5Hz) with the implanted device, was smoothed temporally over the preceding 1.2 seconds (cf. Vansteensel et al., 2016). Subsequently, recording files were concatenated into one long data file and matched to a file with date and time information. Timepoints without brain data were filled with NaN (‘not-a-number’) values. Data was then epoched by taking the average of the preceding 1 minute of data (Ramot et al., 2013) and z-scored by using the mean and standard deviation (SD) of the day data. Statistics were only applied to non-epoched data.

### 2.6 Data Analysis: Comparing Day and Night ECoG Signals

For initial exploration of the LFB and HFB power dynamics, we plotted the normalized, epoched LFB and HFB signals from the first day-night dataset (night data > 6 hours duration with matching daytime dataset) of each month (Table 1). Also, using all non-epoched day-night datasets, we compared power and variance between individual days and subsequent nights (after removing outliers > 3 SDs) by subtracting mean power during the day from mean power during night.

Next, we computed the LFB and HFB power and SD for every available, non-epoched day (499) and night (449) for which a recording file of at least 1 hour duration was available (Table 1). We produced day and night boxplots for the 4-week datasets and used paired-sample t-tests for statistical comparison between day and night data of the 4-week periods. When Shapiro-Wilk normality tests revealed non-normality of a 4-week dataset, the non-parametric Wilcoxon test was used.

### 2.7 Data Analysis: Performance of Daytime BCI Decoder at Night

We investigated how many unintentional click commands and escapes would have occurred during the night, if the decoding algorithms the participant used to produce these BCI commands during daytime at-home use of the BCI, were applied offline to nocturnal data that was acquired in nights in which the participant did not yet use the BCI (initial night measurements, yielding 86.2 hours of data; Table 1). We present an overview of the number of unintentional click-commands and escapes per hour.

### 2.8 Nightmode Decoder Development

#### 2.8.1 Data Collection

We developed a dedicated nightmode decoder to provide the participant with a BCI system she could leave on during sleep with minimal false positive activations, while giving her the option to call a caregiver at night when needed. An additional requirement was that the system should not depend on visual stimuli, because the participant’s eyes were kept closed at night with medical tape, to prevent uncontrolled opening and subsequent drying of the cornea. Data recorded during seventeen nights in the period of Dec 2017 to Jan 2020 (weeks 111-220) were used to investigate the spontaneous occurrence of decoder activations with the aim to develop a nightmode decoder that would produce a minimal number of false positives. Additionally, data recorded during daytime research sessions was used to ensure the participant’s ability to reliably activate the nightmode decoder (i.e., few false negatives). In these research sessions, the participant tested the activation of the nightmode decoder by performing different mental strategies. Several strategies proved unsuccessful. An overview of the attempted strategies is presented in the Supplementary Results and Supplementary Table S1 and S2. Feasibility of the strategy that was eventually implemented and utilized was tested in 14 repetitions of a research task (recorded with the first implanted device in weeks 220-244 across five research sessions; see Table 1).

#### 2.8.2 Nightmode Decoder Solution

The implemented nightmode decoder employed a sequence of correctly timed *‘nightmode events’* that were based on the LFB rebound: a strong LFB increase that occurs upon the termination of attempted movement. To cue the nightmode events, the participant used the timing of her ventilation machine, which was usually set to 15 cycles per minute (*CPM*) and was sometimes adjusted to 14 or 16 CPM. The mental strategy the participant executed was attempted hand movement during one 4-second ventilation cycle (when using 15 CPM), and rest during the next ventilation cycle. This sequence was repeated, leading to a LFB rebound approximately every 8 seconds. An individual nightmode event was registered when the normalized LFB signal passed a threshold of −0.4 for 2 seconds. Normalization was based on a 30-second calibration period that was completed before switching on the nightmode. During the 30-second calibration, the participant attempted three hand movements. The nightmode event interval parameter was set to 6-10 sec (centered around the 8-second duration of two ventilation cycles), allowing for timing deviations and changes in the ventilation settings from 14-16 CPM. Other algorithm parameters were the required number of cycles, the rate, and the variance cutoff (Table S3). The algorithm applied a sliding window to the preceding x seconds, with x being the number of cycles multiplied by the duration of one active-rest cycle (8 sec when using 15 CPM). The number of correctly timed nightmode events in the window was counted. When all requirements were met, the BCI system was activated, enabling the use thereof to call the caregiver (more info in *Nightmode User Interface*).

#### 2.8.3 Nightmode User Interface

The participant was able to set the system to enter nightmode with regular click commands. Alternatively, a caregiver could switch on the nightmode by using the touch screen. Immediately after the 30-second calibration and switching on the nightmode, the participant often performed a test BCI activation, and repeated the 30-second calibration if the test was unsuccessful. Supplementary Figure S2 and S3 show all user interface screens of the nightmode and its logic.

### 2.9 Data Analysis: Nightmode Performance

We present information on frequency of at-home use and performance metrics of the nightmode. Performance data was collected by caregivers on night-duty, who were asked to assess whether a BCI activation at night was intentional or not (true/false positive), and to ask in the morning whether the participant tried to activate the system and if that was successful (true /false negative). Finally, based on qualitative reports, we present several real-life examples of reasons for using the nightmode to call a caregiver, and user satisfaction.

### 2.10 Data Availability

Data can be shared by the corresponding author upon reasonable request.

## 3. RESULTS

### 3.1 Comparing Day and Night ECoG Signals

#### 3.1.1 Initial exploration

Exploring LFB and HFB power during the day and night revealed that both power and variance often appeared larger in night data than in the day data (Figure 1). Indeed, mean power was higher in 77% (161/209) and 60% (125/209) of nights for LFB and HFB power respectively, and likewise variance of power was higher in 66% (138/209) and 76% (159/209) of nights respectively (Figure 2).

**Figure 1:**
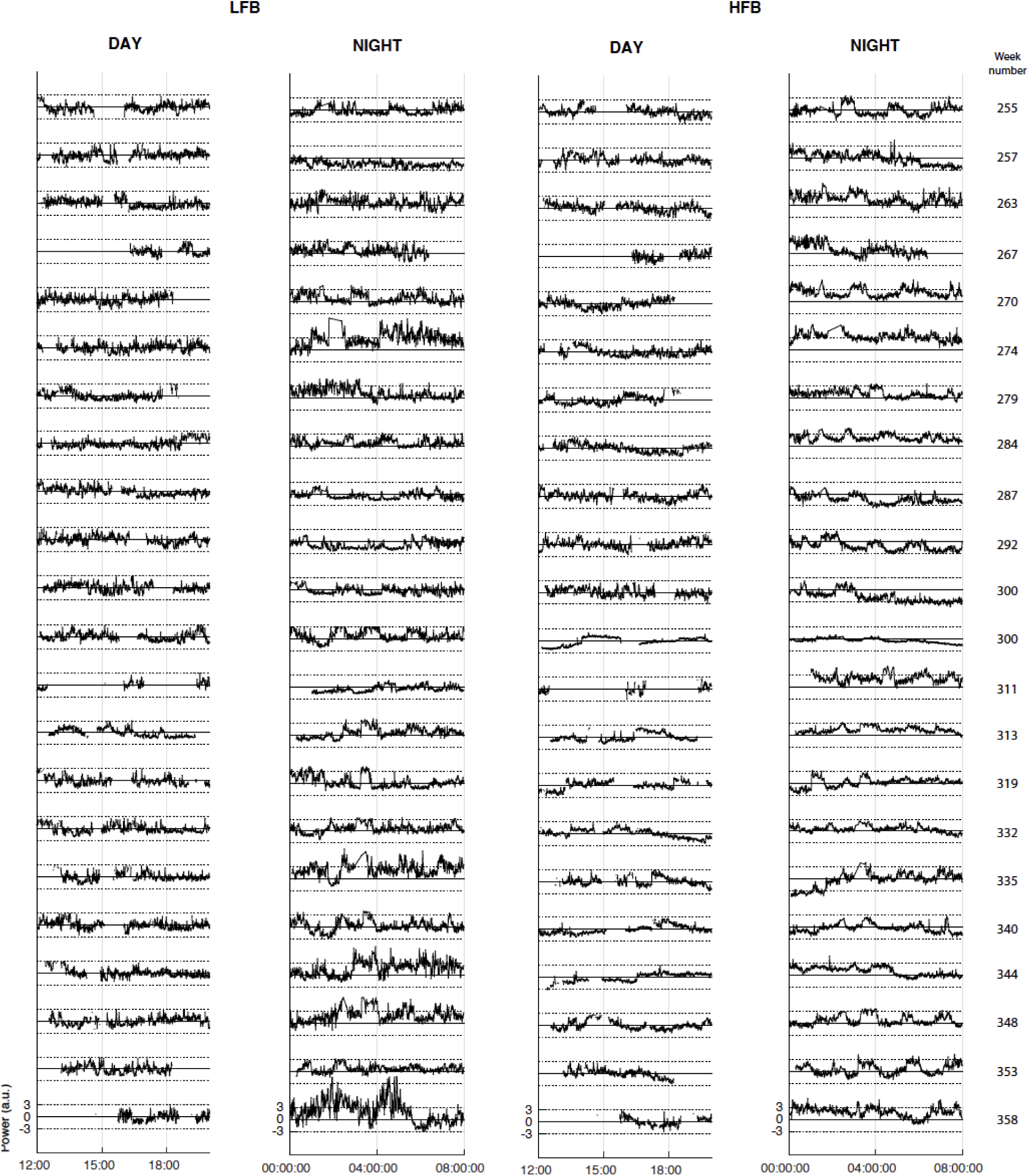
Epoched day and night data (LFB and HFB). Data from the first day-night dataset of each month (weeks after implantation indicated on the right side), normalized based on the mean and SD of the day. This is based on night data of at least 6 hours that followed a day dataset. As a result, mean and SD of day data was always 0 and 1. Y-axes units are arbitrary power units. For reference, each graph has horizontal lines at 0 (thick) and at −3 and 3 times SD (dotted). The first available day-night dataset from July 2021 was recorded at the end of the month (due to an incorrect Windows date and time in the preceding period), explaining why two datasets are plotted for week 300.

**Figure 2:**
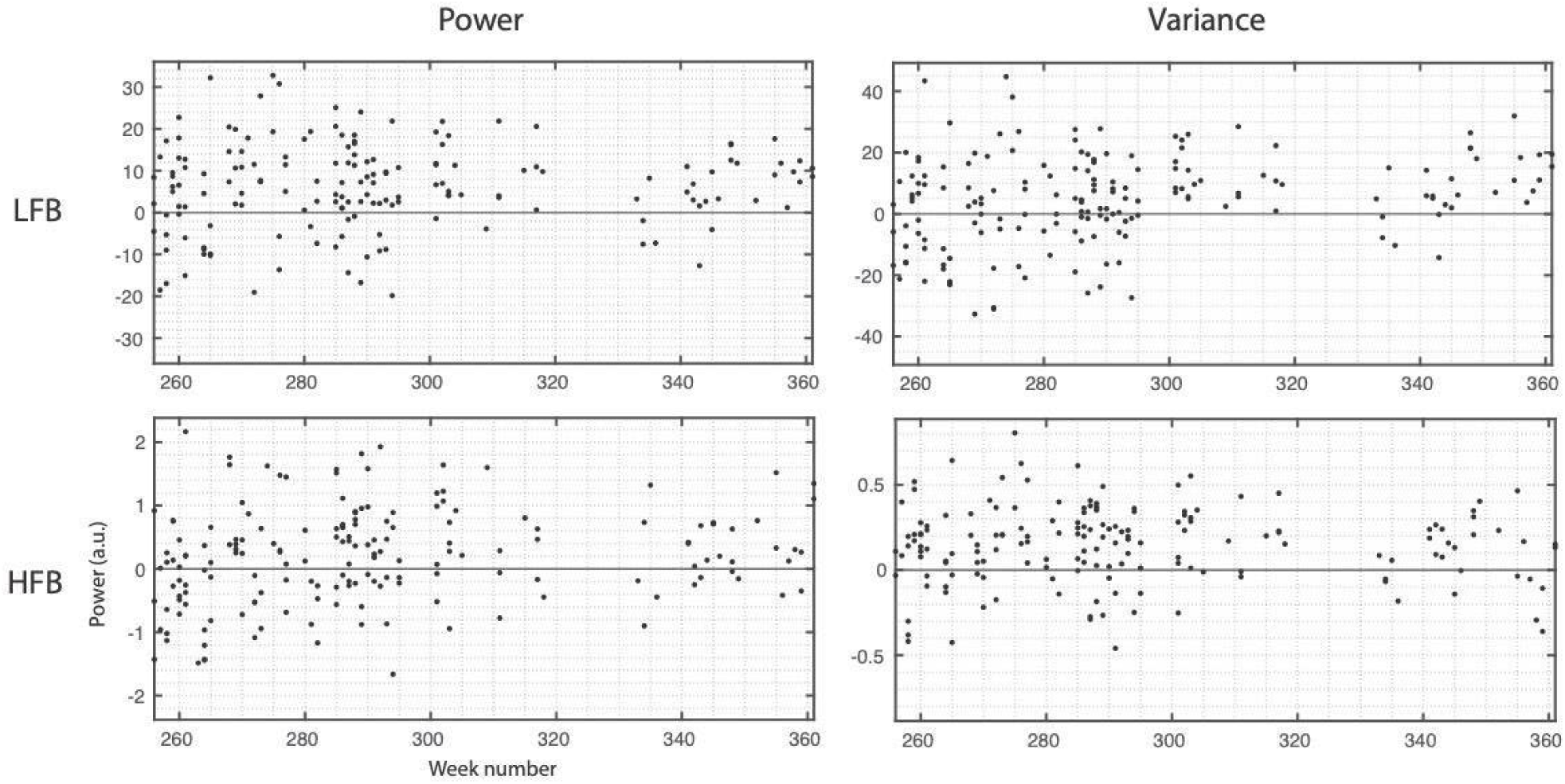
Day-night comparison from non-epoched data. Day metrics (mean power and variance) were subtracted from the respective night metrics. Y-axes indicate arbitrary power units. For visual reference, 0 is centered and indicated by a horizontal line. Datapoints above 0 indicate that the metric was higher at night. Data is plotted chronologically with weeks since implantation of the ECoG electrodes on the x-axis.

#### 3.1.2 Statistical Analysis

Statistical analysis of the 4-week datasets indicated that LFB and HFB power were significantly higher at night than during the day in 58% of the 24 4-week periods (p < 0.05; Figure 3; analysis based on data presented in Supplementary Figure S1). In only one 4-week period, LFB power was significantly lower at night (p < 0.05). For the other periods, there was no significant difference between day and night for both LFB and HFB power. Variance was significantly higher at night for LFB and HFB in 54% and 88% of periods, respectively. Only one 4-week period (the same period as for LFB power) showed significantly lower power variance during the night for LFB and HFB (p < 0.05). Other periods did not differ significantly. Note the steady decline in both power and variance over time in Figure 3, which we attribute to ALS progression (Vansteensel et al., 2024).

**Figure 3:**
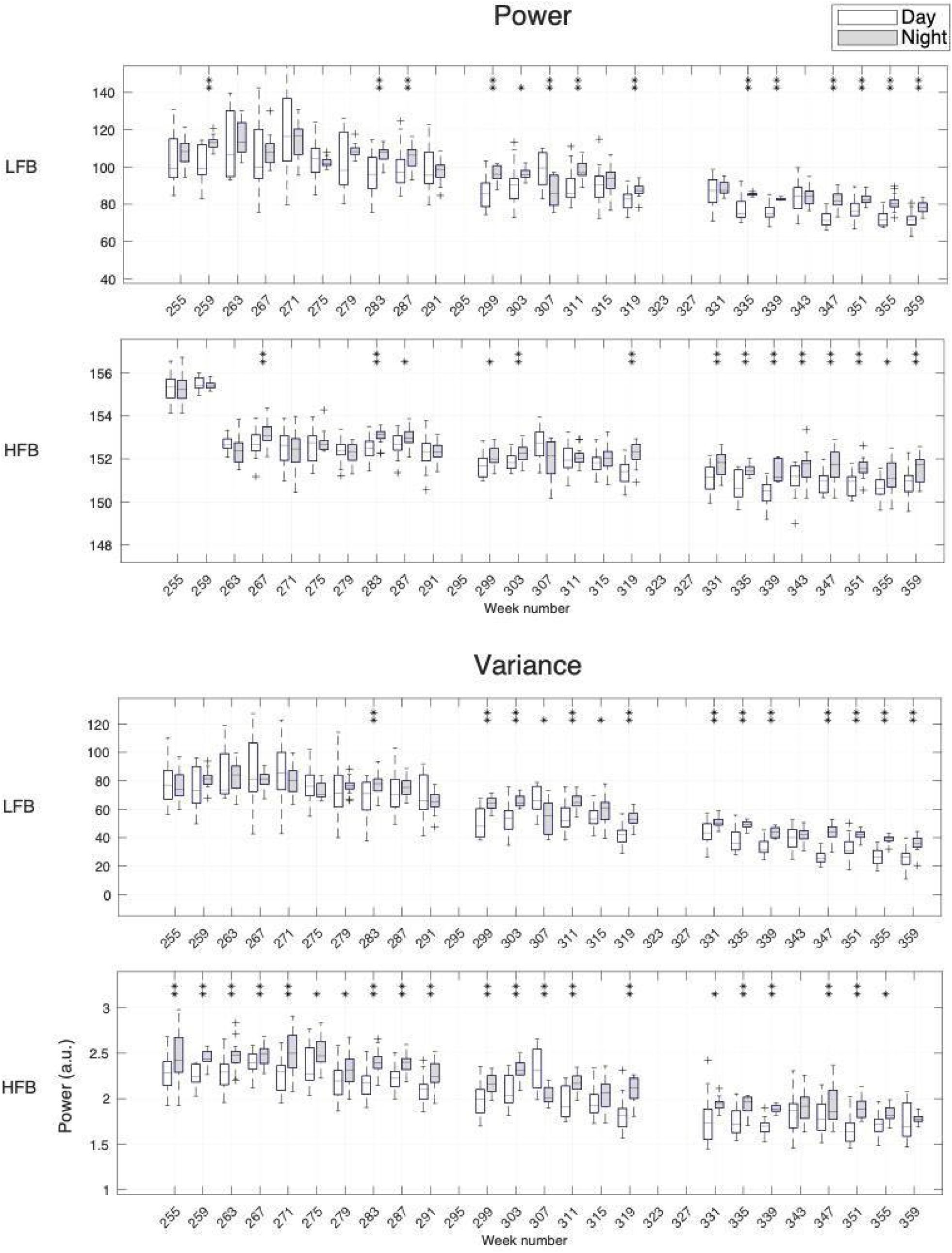
LFB and HFB power (top panels) and variance of power (bottom panels) during day (white) and night (gray). Power and variance were computed for each day and night and grouped into boxplots for each 4-week dataset (median, interquartile range and min/max values). Y-axes indicate power in arbitrary units. Week numbers indicate the start of each 4-week period. White boxplots indicate day data and gray boxplots indicate night data. A sudden drop in HFB power was observed at week number 263. No explanation was found for this. HFB variance was not affected by this drop, nor was LFB power or variance. Asterisks indicate statistically significant differences between day and night (* p < 0.05; ** p < 0.01).

### 3.2 Performance of Daytime BCI Decoder at Night

Offline application of the daytime decoders for brain-clicks and escapes on data that was acquired during the initial, passive, night measurements in a period where the BCI was not yet used during the night, resulted in 245 unintentional clicks and 13 unintentional escapes per hour during the night (Figure 4).

**Figure 4:**
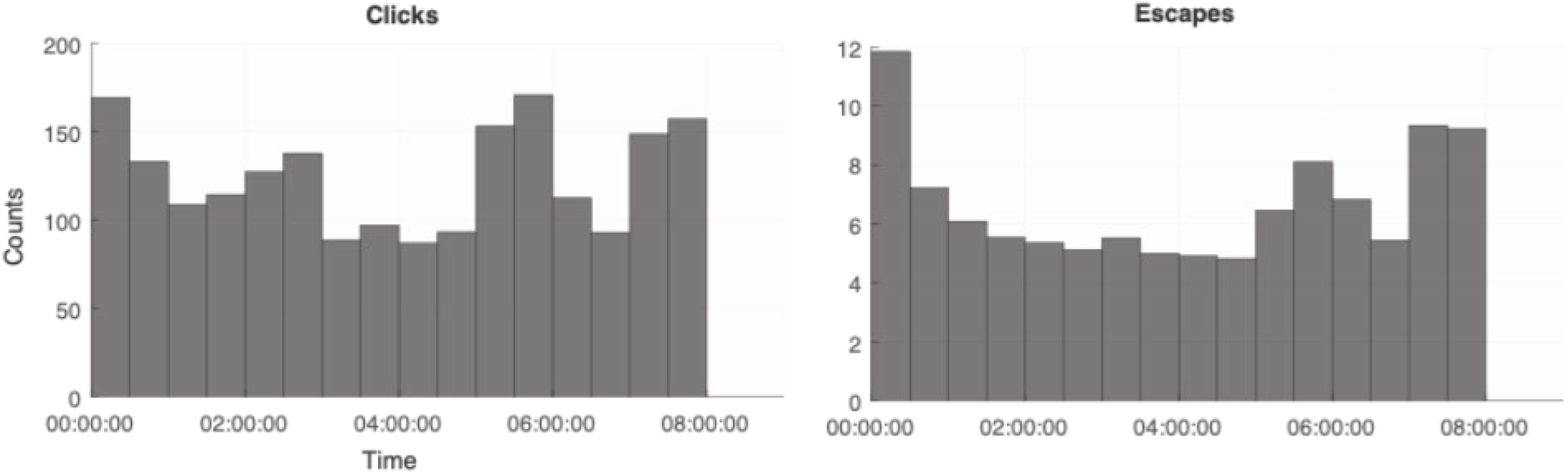
Distribution of unintentional clicks and escapes. The number of unintentional clicks (left) and escapes (right) plotted in 30-minute blocks, calculated on the initial night measurements performed before installing the nightmode (86.2 hours of data recorded across 17 nights). Each bar indicates 30 minutes of the period between 0:00h - 8:00h, which was considered night, as per caregiver information.

### 3.3. Nightmode Decoder and Performance

#### 3.3.1 Nightmode Parameters

The parameters for the nightmode decoder that was implemented and utilized are shown in Table S3. With these parameters, the system was activated as intended (true positive) in 13 out of 14 research task repetitions acquired during research sessions (Table 1) and no false positives were present in the initial night measurements. The only parameter that was adjusted in the period the participant used the nightmode at home was the number of cycles. An example of the pattern of signal changes required to activate the system and call the caregiver at night, recorded during a research session, is shown in Figure 5.

**Figure 5:**
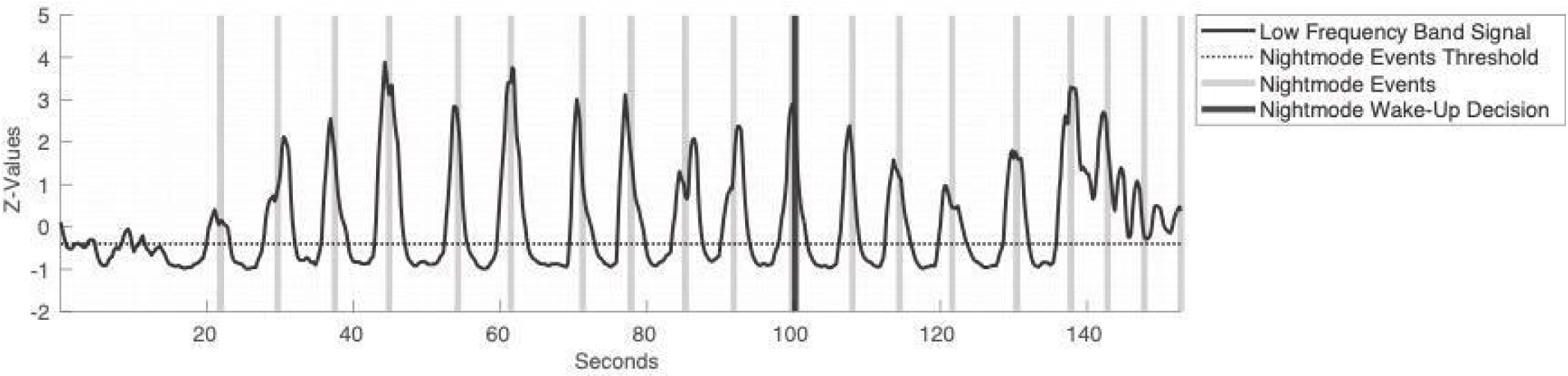
Nightmode sequence example. LFB power recorded during a research task in which the participant was asked to follow the block design of her ventilation machine (4 seconds of attempted hand movement, 4 seconds of rest). Besides an auditory start (at 20 seconds) and stop cue (at 140 seconds) no other task cues were provided. In this case, the nightmode decoder was activated after 11 nightmode events even though it was set to 12 cycles. This happened because the rate was set to 0.01, allowing for one false negative at any point in the sequence. The absence of any false negative nightmode event between cycles effectively activated the nightmode decoder one cycle sooner.

#### 3.3.2 Nightmode Use and Performance

Between September 2020 and October 2022, the participant used the BCI in 494 nights. Between week 255 and week 334, nightmode performance was logged for 337 nights (Figure 6A). The number of logged nights decreased in the final months, when overall use of the system became more erratic and communication at night became more difficult (Vansteensel et al., 2024). As a result, caregivers could no longer determine if a nocturnal caregiver call was intended. Caregivers stopped filling out the performance logs from April 2022 onward. Nevertheless, the nightmode continued to be in use on the participant’s request until early October 2022 (week 361). In consultation with the participant and her caregivers it was then decided to no longer use the nightmode, due to unreliable performance and communication.

**Figure 6:**
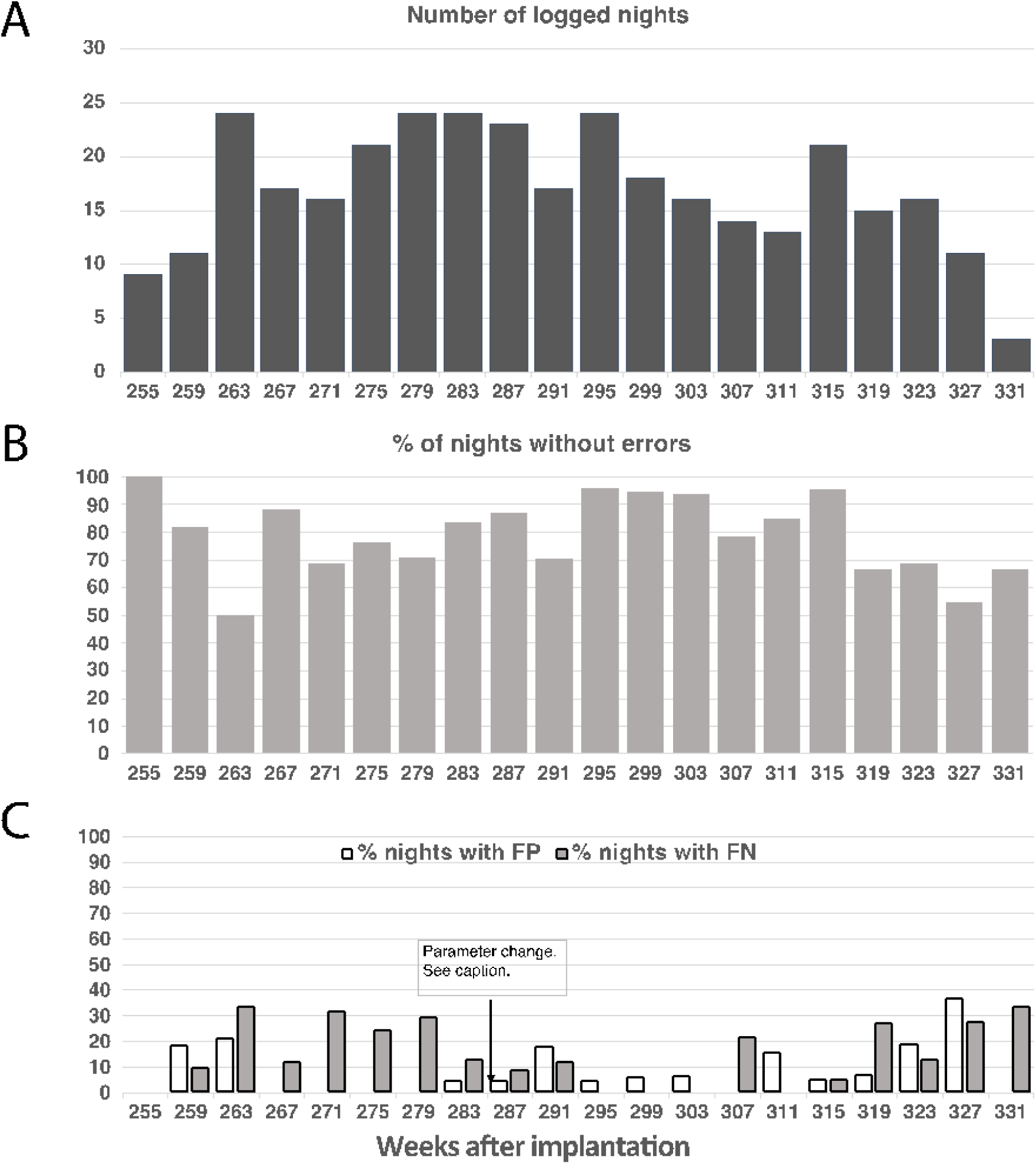
Nightmode data overview and performance. X-axes indicate week number since electrode implantation, with each week number representing the start of a 4 week period. A: Number of nights per 4-week period for which performance was logged. B: The percentage of nights without any false positives (FP) or false negatives (FN). C: The percentage of nights that contained 1 or more false positive (FP) and 1 or more false negatives (FN). Notably, the ‘number of cycles’ parameter (see Table S3) was changed in week 284 from 12 to 11. In week 286 it was changed from 11 to 10, after which two false positives occurred in the subsequent night. The number of cycles was then reverted to 11. This parameter change led to 9% and 14% of nights with FPs and FNs, respectively in the period from April 2021 until March 2022 (week 283 – week 334, when the last nights were logged).

The nightmode functioned without errors in 79% of all logged nights (Figure 6B). In 33% of all logged nights, only true negatives occurred (i.e., no nocturnal caregiver calls were attempted and no false positive occurred). 58% of nights contained true positive nocturnal caregiver calls and on average there were 2.3 caregiver calls in those nights. 15% of all nights contained false negatives (on average 2.4 in those nights) and false positive caregiver calls occurred in 8% of nights (once every ± 12 nights; Figure 6C). Individual nights could contain combinations of false and true, and positive and negative events.

Based on qualitative reports kept by caregivers, the participant used the nightmode to call caregivers mostly for care-related requests, such as lung suctioning or medication. The participant reported satisfaction with the nightmode on multiple occasions to researchers during research sessions.

## 4. DISCUSSION

For real-life use, a BCI communication device should be available and function reliably 24/7. We show, in a person with late-stage ALS who had an implanted BCI system, that decoders that performed well during daytime generated high rates of false positive detections of BCI actions during sleep, related to different nocturnal signal characteristics, which in effect would render the system useless at night if not accounted for. We also present a nightmode decoder solution that allowed the user to activate the BCI system and call caregivers reliably at night, with few false positive activations.

### 4.1 Comparing Day and Night ECoG Signals

The most noticeable observations when comparing day and night signals were that LFB (10-30Hz) and HFB (65-95Hz) power and variance were often higher during the night than during the day, whereas the opposite occurred only sporadically. This result contradicts that of (Cantero et al., 2004), who found higher gamma variance during wakefulness than during sleep in four able-bodied epilepsy patients. This discrepancy may be explained by the motor impairment of the participant of this study, and the associated lack of activation of the sensorimotor cortex. In the period of study (i.e., after week 255 post-implantation), the participant only moved when communicating yes-no with the corner of her mouth. In addition, cortical activity associated with BCI use and passive movement of limbs can be expected to occur far less frequently than the almost continuous activation expected due to movement and sensory activity in able-bodied people, even when they are bedridden such as in (Cantero et al., 2004).

The nature of the variability in HFB and LFB power and variance at night, within and between the 4-week periods, is currently unclear. One potential explanation for the variability is that the participant sometimes napped during the day. Naps of more than 30 minutes long have been suggested to affect nighttime sleep (Yoon et al., 2003). No logs on naps or actual nocturnal sleep were kept (due to caregiver workload constraints). Second, it cannot be excluded that different levels of passive movements, such as those related to caregiving, account for differences of LFB and HFB power across different day/night combinations. Third, different sleep-stages (NREM/REM) are known to be associated with different neural signal characteristics (Abe et al., 2008; Matarazzo et al., 2011; Šušmáková, 2004). Whereas the current dataset did not allow to distinguish between sleep-stages, different distributions thereof between days/weeks/months may be associated with different power and variance. Finally, research has shown that sleep is often affected in people with ALS (Lucia et al., 2021; Zhang et al., 2023). Specifically, sleep efficiency is often decreased and wake time after sleep is increased. In our study, differences in sleep quantity and quality between nights may have led to variability in HFB and LFB power and variance. Further research will be needed to gain a full understanding of the effects of sleep and any ALS-related changes therein, on BCI control signals in the sensorimotor cortex.

### 4.2 Performance of Daytime BCI Decoder at Night

Unintended BCI classifier activations should be kept to a minimum at all times but especially during the night, since they could interrupt the sleep of both the user and caregivers. We found that many false positive events were produced when classifier parameters optimized for daytime usage were applied to night data. The data used for this offline analysis was collected during the period in which the participant was not yet using the BCI at night. Therefore, all clicks and escapes that were detected by applying daytime decoders on nocturnal data can be considered unintentional, false positive BCI activations. Importantly, whereas BCI performance in this participant eventually declined due to ALS progression (Vansteensel et al., 2024), daytime decoders were highly accurate in the period the data for this assessment was acquired (∼year 2-4 after implantation; c.f. Figure 1C of Vansteensel et al., 2024). We therefore conclude that the erroneous decoding of clicks and escapes during nighttime was not caused simply by a lack of parameter optimization, but by spontaneous changes in the nocturnal signals for which the daytime decoder was not optimized..

### 4.3 Nightmode Decoder

The nightmode decoder was used by the participant for over two years. It allowed her to call caregivers when needing attention at night, The relatively long decision period of the nightmode algorithm (approximately 1.5 minutes) seems impractical at first glance, but was considered usable by the participant, particularly given the absence of any other option. The relative success of this nightmode underscores the importance of applying an iterative and user-centered design approach in these types of clinical trials.

The impact of daytime decoding errors on daily use of the BCI system is limited, provided they occur sporadically. At night on the other hand, a single false positive caregiver call can negatively affect sleep quality of the user and of the caregivers. The performance requirements for BCI use at night can therefore be considered more strict than those for daytime usage. For example, up to December 2021 (week 318) 1 in 12 nights contained an unintended caregiver call, which was deemed acceptable by the participant and her care team. However, when the number of false positives increased (e.g. after a parameter change in April 2021 to 2 false positives in one night) or when it was no longer possible to ascertain the validity of the system, caregivers were reluctant to rely on the system. A second requirement is the need for unobtrusive cues instead of visual or auditory cues that may disrupt sleep or that may be otherwise unusable for particular individuals. The current solution relied on the pace of the ventilation machine, which turned out to work well for the participant. Similar methods of using unobtrusive timing cues for generating precisely timed changes in neural signals that are unlikely to occur spontaneously during sleep may work for other users of similar BCI systems, but parameters will most likely require customization.

### 4.4 Future Directions

The current BCI system provided a one-dimensional control signal. The fact that the participant required at least 11 active-rest cycles to minimize false positives at night underscores the large variability of sensorimotor brain signals. Other ECoG-based BCI systems that employ multidimensional control signals (e.g. (Anumanchipalli et al., 2019; Metzger et al., 2023; Moses et al., 2021) or BCI systems using intracranial micro-electrode arrays (e.g. (Pandarinath et al., 2017; Stavisky et al., 2018; Willett et al., 2023, 2021) are being tested, but to our knowledge, are not used yet by the users in real-life without researcher presence. It is likely that also these more advanced BCI systems, if they were to be implemented in the daily life of the people they are meant to serve, will have to be adjusted to cope with circadian or sleep-related signal changes in the neural signals. Indeed, (Rubin et al., 2022) reported false positive decoded events (2-D cursor movements) in data recorded during sleep in a BCI participant with a multi-electrode array in the sensorimotor cortex.

In the nightmode, activating the system was accomplished through an intentional mental strategy. An automatic classifier that detects when someone is asleep or awake would be preferable as it does not require any active input from the user, but this requires reliable, automatic sleep-stage detection from intracranial signals, which the current dataset was unable to provide. Such feature requires more research, and may benefit from recordings of brain regions other than those used for direct BCI control.

## 5. CONCLUSIONS

We report on day-night differences in ECoG signals measured in an implanted BCI user with late-stage ALS. We show that LFB and HFB sensorimotor signals used for BCI control were subject to spontaneous signal perturbations during the night, and that applying BCI decoder parameters optimized for daytime use to night data caused many unintended decoder activations. We present for the first time a nightmode decoder solution that allowed a BCI user to reliably call a caregiver at night. Providing a BCI system that can cope with sleep-related brain signal changes is the difference between a BCI system that is available 24/7 and a BCI system that is usable 24/7.

## ACKNOWLEDGEMENTS

The authors thank the participant of this study, and her family and caregiver team, whose courage, dedication, insights, and hospitality made this research possible. Thanks to Dr. Joram van Rheede for useful discussions about 24/7 neural signals.

