## Supplementary material for "DO NOT LOSE SLEEP OVER IT: IMPLANTED BRAIN-COMPUTER INTERFACE FUNCTIONALITY DURING NIGHTTIME IN LATE-STAGE AMYOTROPHIC LATERAL SCLEROSIS": SupplemetaryMaterials

### Figure S1: Data overview

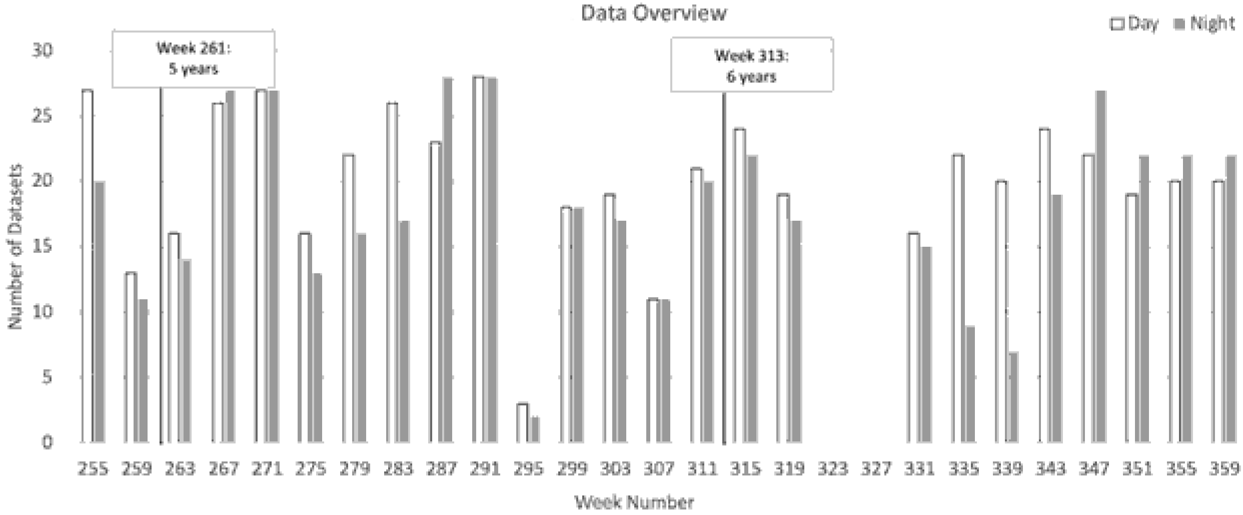


***Figure S1: Data overview.*** *Data was recorded with the implanted device between September 2020 until October 2022. The x-axis shows weeks since implantation, with week numbers indicating the first week of each 4-week period. The graph indicates for how many days and nights data was available (recording files of at least 1 hour duration) in each 4-week period ('4-week datasets' in Table 1 of the main paper). Some data recorded in July 2021 (around week 295) and January-February 2022 (around week 323-327) could not be used, because the Windows date and time information of the home-use tablet (used for timestamping brain data) was incorrect. Even though some data was recorded in the 4-week period starting on week 295, it was not analyzed statistically.*
*We have indicated the 5- and 6-year marks after implantation, for convenience and to facilitate comparison with other manuscripts with data from this participant.*

#### Figure S2: Escape Window

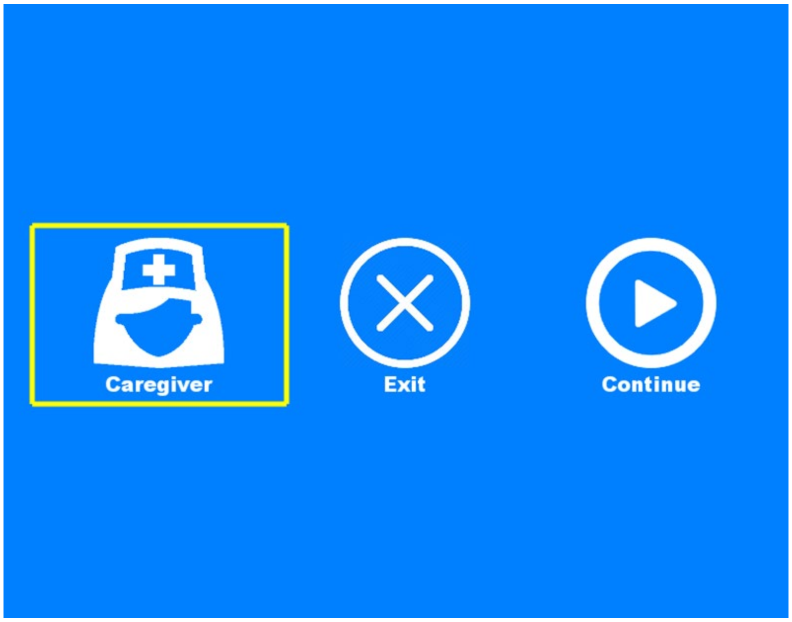


***Figure S2: The escape window***. *This window replaced the display of any previously active application when a daytime escape command was issued by the participant and detected by the system. The user could then select one of the three options with regular clicks as these were sequentially highlighted in switch scanning mode. The caregiver option played a buzzer sound; the exit button exited the currently active application (e.g., commercial spelling software) and returned the user to the main menu of the custom BCI software; continue closed the escape window and returned to the previously active application. The escape was also linked to a standby mode that the user could activate (using regular clicks). During standby, the BCI system did not respond to regular clicks but only to escape events, making it a useful feature for activities that required daytime BCI availability but not active usage, such as watching tv or during a walk.*

### Figure S3: Nightmode User Interface

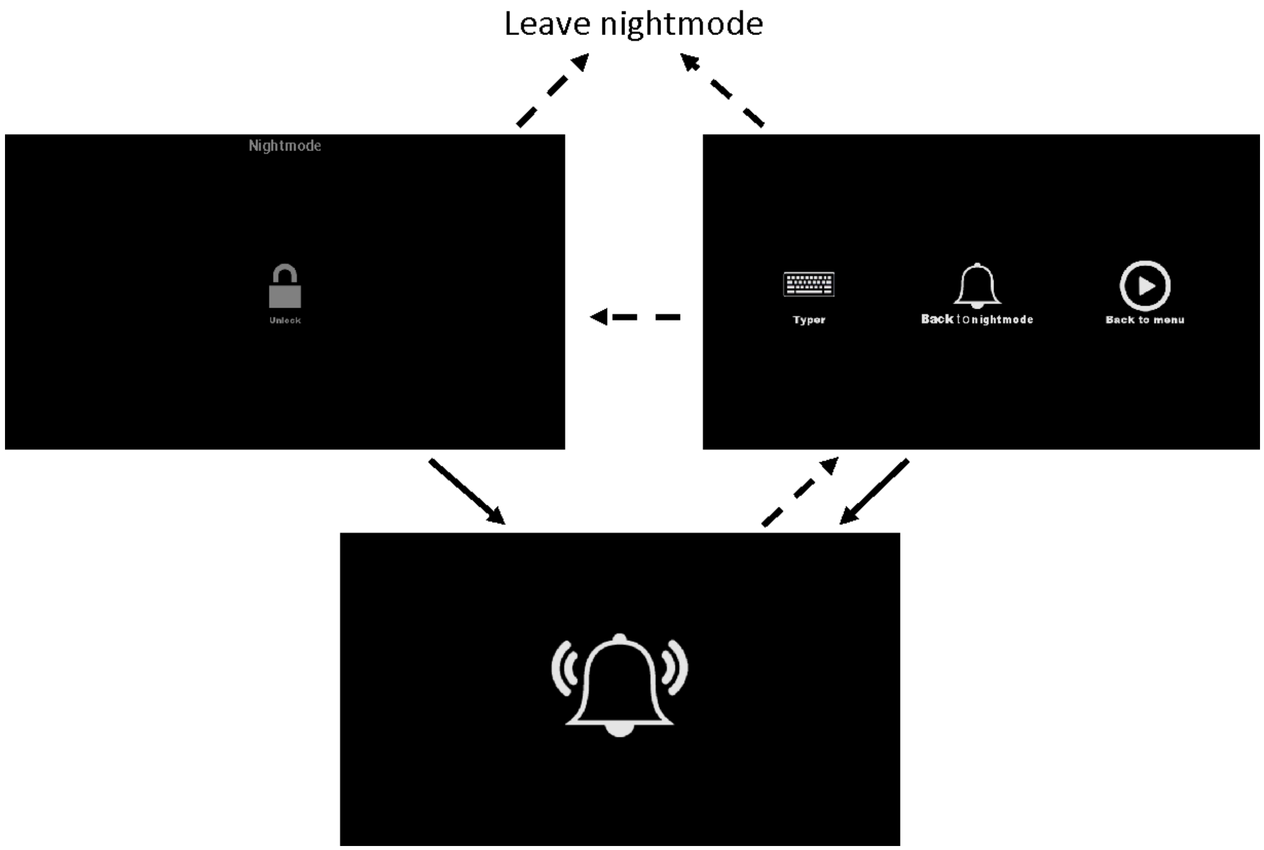


***Figure S3: User interface of the nightmode.*** *Solid lines indicate actions the participant could make, and dashed lines indicate actions caregivers could make using the touch screen. The screen brightness was set to 0% during nightmode (top left). When the user activated the system using the nightmode decoder, the tablet-computer produced a buzzer sound (bottom) until a caregiver touched the screen (top right). To prevent the participant from accidentally leaving the nightmode, only caregivers were able to leave the nightmode using the touch screen (using the 'typer' or 'back to menu' buttons). In case caregivers forgot to return the system to nightmode (top left) after a caregiver call, the participant retained the ability to activate the system (solid line from top-right panel to bottom panel), although the screen brightness would not be 0%.*

### Supplementary Results

#### Nightmode: Strategies

We tested several nightmode strategies offline. First, we attempted using existing filters with adjusted parameters and visual cues (Table S1).

| **Method** | **Signals Used** | **Mental Strategy** | **Classifier Info** | **Electrode Pair(s)** | **Result** |
|---|---|---|---|---|---|
| Regular clicks | Sensorimotor control signal (HFB – LFB power) | Attempted finger movement | Daytime click parameters (1 sec above threshold) | E2E3 | Many false positive activations at night |
| Regular escape | Sensorimotor control signal (HFB – LFB power) | Sustained attempted finger movement | Daytime escape parameters (>7.6 sec above threshold) | E2E3 | Many false positive activations at night |
| Longer escapes | Sensorimotor control signal (HFB – LFB power) | Sustained attempted finger movement | Escape duration up to 1 minute. Other escape parameters unchanged. | E2E3 | Many false positive activations at night |
| Longer escapes with signal adaptation to account for the potential effect of slow, large power fluctuations observed at night. | Sensorimotor control signal (HFB – LFB power) | Sustained attempted finger movement | Adaptation based on a sliding window of duration 1-5 minutes.<br>Long escape duration parameters of 60 second were used. | E2E3 | Many false positive activations at night |
| Sequence of sensorimotor activity and rest with varying durations, tested in research | Sensorimotor control signal (HFB – LFB power) | Cued attempted finger movement and rest | Sequences of active (above threshold) and rest (below threshold) activity. Active periods were varied (3, 5, 7, or 9 seconds) and rest periods were 3 seconds | E2E3 | Many false positive activations at night and false negatives during |

| | | | | | |
|---|---|---|---|---|---|
| sessions with visual cues. | | | (chosen based on time required for control signal to dip below threshold (Leinders et al., 2017) ). Up to three sequential trains of active and rest were tested. | | research sessions. |
| Sequential motor and working memory activity | Sensorimotor control signal (HFB – LFB power). DLPFC: HFB power. | Comprised three mental strategies: Attempted finger movement; mental arithmetic; rest. Trial length of each strategy was 6 seconds, with a 6 sec rest period (R) between motor (M) and working memory (WM) trials. Trial layout: M-R-WM-R-M-R-WM (4 active trials and 3 rest trials) | Classifier based on sequential increases in activity on sensorimotor electrodes (by attempted hand movement) and on DLPFC electrodes (by mental arithmetic). | E2E3 & E9E11 (DLPFC)* | No false positive activations at night, but false negatives in research sessions. DLPFC signals were unavailable in the second implant. |
| * E9E11 (when connected to the DLPFC strip in the first implanted device) was recorded in some early night recordings that were used for testing | | | | | |

***Table S1. Attempted Nightmode Methods 1.*** *Initial strategies based on the existing click and escape filters, and on brain signals elicited by having the participant follow visual cues, did not reach sufficient performance.*

The aforementioned strategies led to unacceptable rates of false positives or false negatives. Next, in consultation with the participant, we tried longer patterns that required signal modulation for more than one minute. Moreover, because she could no longer open or close her eyes at night, and because auditory stimuli would affect sleep quality, the solution had to be independent of visual and auditory cues. We therefore tested if the participant was able to use the invasive positive-pressure ventilation machine as a timing cue. The participant's ventilation machine was usually set to 15 cycles per minute (*CPM*). We asked the participant to attempt right-hand finger movement during one 4-sec ventilation cycle (i.e., one inhale-exhale sequence) and relax during the next cycle, and we measured her brain signal as she executed this strategy for two minutes. We recorded 14 of these datasets during daytime research sessions to ascertain whether the participant could follow this block-design pattern with her brain signal and to find decoders with few nighttime false positives and daytime false negatives. Classifiers applied to the sensorimotor control signal elicited upon following this paradigm included a simple threshold-based match filter, a power-transform filter, and a state filter that classified states (active or rest) from the signal (Table S2). These filters did not reach the desired performance. However, the paradigm of following the ventilation machine as a pacer eventually led to the implemented night-mode solution using only the LFB power signal. That paradigm is summarized in Table S2 and detailed in the main text, section *Methods: Nightmode Decoder Solution*.

| **Method** | **Signals Used** | **Mental Strategy** | **Classifier Info** | **Electrode Pair(s)** | **Result** |
|---|---|---|---|---|---|
| Repeated sequence of sensorimotor activity and rest | Sensorimotor control signal (HFB – LFB power) | Cued attempted finger movement and rest. Sequences of active (above threshold) and rest (below threshold) activity, with duration of active and rest based on the ventilation machine of the participant (4 seconds). | 3 different classifiers were tried: | E2E3 | |
| | | | 1) 4 - 8 cycles of active and rest (32 - 64 second in total); at least 80% match between above and below threshold of brain signal and the block design was required. | | 1) Many false negatives in research sessions. |
| | | | 2) Applied power transform to window sliding to most recent 30 second from control signal, with power transform center frequency centered around 0.125 Hz (the frequency at which we expect most power with an 8 sec cycle). When power passed a threshold (values ranged from | | 2) Many false positives in night data or -with other parameters settings- false negatives in research sessions. |

| | | | | | |
|---|---|---|---|---|---|
| | | | 1-20) for x seconds (duration ranged from 30 -90 seconds), an activation was registered. | | |
| | | | 3) State filter that detects active and rest blocks from brain signal (after x seconds above or below threshold, respectively). Required block duration of each condition was tested for different minimum and maximum durations (2-8 sec). Active-rest alternations were tested up to a total duration of 2 minutes. | | 3) Many false positives in night data or - with other parameter settings - false negatives in research sessions. |
| **Eventually used paradigm** | | | | | |
| **Method** | **Signals Used** | **Mental Strategy** | **Classifier Info** | **Electrode Pair(s)** | **Result** |
| Repeated sequence of sensorimotor activity and rest | Sensorimotor LFB only | Cued attempted finger movement and rest. Sequences of active (above threshold) and rest (below threshold) activity, with duration of active and rest based on the ventilation machine of the participant (4 seconds). | LFB power was converted into 'nightmode events', using the same algorithms as those used for regular clicks. Nightmode events were expected to occur during LFB rebounds (which occurred after cessation of movement attempt) every ± 8 sec. When a sequence of 12 nightmode events was correctly timed (6-10 sec between nightmode events) and when additional requirements were met, the system would be activated. | E2E3 | Successful |

***Table S2. Attempted Nightmode Methods 2.*** *These methods were based on new algorithms and cues from the ventilator machine and using longer (> 1 minute) patterns.*

| Nightmode parameters | | |
|---|---|---|
| Click interval | 6 – 10 sec | Always 6 – 10 sec based on ventilation parameters |
| Number of cycles | 12 | Defined the number of required nightmode events. |
| Rate | 0.01 | Defined how many false negative/false positive events errors were allowed. The rate was multiplied with the number of cycles and then rounded up to the next integer. For example, using a rate of 0.01 and number of cycles of 12, one false negative and one false positive were allowed (0.01 * 12 = 0.12., rounded up to the next integer → 1). |
| Variance Cutoff | 0.8 | Defined the minimum required standard deviation of the LFB signal in the sliding window. If the SD was lower, no system activation and nocturnal caregiver call signal would occur. |

***Table S3: Initial nightmode parameters.*** *LFB rebound signals occurring after cessation of attempted movement were used as the signal source for nightmode events. Nightmode events were produced when the LFB signal passed -0.4 for two seconds. For a nightmode system activation and caregiver call signal to be triggered, two requirements had to be met: the correctly timed number of clicks had to exceed the rate multiplied by the number of cycles, and the variance in the sliding window had to exceed the variance cutoff.*
